# Childhood Correlates of Genetic Propensity to Personality Traits

**DOI:** 10.64898/2026.09.10.26362760

**Authors:** Margaret L. Clapp Sullivan, Alex P. Miller, Emma C. Johnson, Nicole R. Karcher, Sarah E. Paul, Joshua J. Jackson, Ryan Bogdan, Arpana Agrawal

## Abstract

The Big 5 personality traits (extraversion, agreeableness, conscientiousness, neuroticism, openness to experience) are moderately heritable correlates of behavior, health, and life outcomes. Characterizing childhood expressions of genetic variation associated with adult personality may inform our understanding of personality development and how personality becomes associated with behaviors and environments. Our phenome-wide association study (PheWAS) of Big 5 polygenic indices (PGIs) among children (n_European-like ancestry_=5,556; n_African-like ancestry_=1,584) in the Adolescent Brain Cognitive Development Study^SM^ (ABCD Study®) across ages 9-11 and 11-13 revealed stable associations between neuroticism PGI and measures of child mental health, openness PGI and cognition, agreeableness PGI and externalizing behaviors, conscientiousness PGI and perseverance, and extraversion PGI and impulsivity. Longitudinal analyses indicated stronger personality PGI associations with some phenotypes (e.g., prosocial behavior) at ages 11-13. Behavioral correspondence to genetic propensity to adult personality is evident in early adolescence while certain correlates of personality PGIs may only manifest later in development.

## Introduction

The Big 5 personality traits (extraversion, agreeableness, conscientiousness, neuroticism, openness to experience) are moderately heritable patterns of thinking, feeling, and behaving that predict critical life outcomes (e.g., educational attainment, health, longevity) as robustly as socioeconomic status and cognitive ability (Beck & Jackson, 2022; Briley & Tucker-Drob, 2014; Graham et al., 2017; Roberts et al., 2007; Schwaba et al., 2025, 2026). Within-individual personality can be highly variable during childhood and adolescence before stabilizing in young adulthood (Bleidorn et al., 2022; Hampson & Goldberg, 2006; Roberts et al., 2006; Wright & Jackson, 2023). Importantly, individual differences in the Big 5 traits in childhood and adolescence are linked to key outcomes at older ages (Wright & Jackson, 2022). For example, children with higher levels of extraversion were more likely to develop alcohol and drug dependence later in life (Newton-Howes et al., 2015). Characterizing the correlates of adult personality in childhood may identify its earliest forms of expression as well as factors (e.g., experiences, behaviors) that could be leveraged to encourage the development of long-term positive social, health, and academic behaviors.

Personality traits in childhood and adolescence differ from adult personality traits in their content and inter-trait correlations (Soto & Tackett, 2015). For example, agreeableness and conscientiousness are more strongly correlated in childhood than adulthood, and facets of extraversion (e.g., sociability, assertiveness) are more weakly intercorrelated in childhood than in young adulthood (Soto et al., 2008; Soto & John, 2014). Shifts in personality also vary by trait; through adolescence and young adulthood, agreeableness and conscientiousness generally increase, while neuroticism tends to decrease through late adolescence into older adulthood (Bleidorn, 2015; Roberts & Mroczek, 2008; Specht et al., 2014). These shifts may occur as a result of both environmental and genetic influences, where trait expression may increase because of continuous exposure to stable environments (e.g., schooling; Brandt et al., 2019) and/or changing impacts of genetic influences (Briley & Tucker-Drob, 2014). Twin and family studies find that genetic variation explains about 30-60% of the variation in personality traits (Polderman et al., 2015; Vukasović & Bratko, 2015), and that the influence of genetic variation on personality shifts across age, with personality being more heritable in early childhood and declining through late adolescence and early adulthood (Briley & Tucker-Drob, 2014; Kandler, 2012; Tucker-Drob & Briley, 2019).

Building upon twin research documenting the moderate heritability of the Big 5 personality traits and initial genome-wide association studies (GWASs) (de Moor et al., 2012; Gupta et al., 2024; Lo et al., 2017), a recent GWAS of personality in 611,037 to 1.14 million adults characterizes the polygenic architecture associated with extraversion, agreeableness, conscientiousness, neuroticism, and openness (Schwaba et al., 2026). This work demonstrates that the Big 5 domains are highly polygenic (i.e., influenced by many genetic variants each with very small effects; Visscher et al., 2021) and genetically correlated with a wide variety of traits, including educational attainment, mental health, and physical health. Although there is evidence that genetic variants associated with personality are consistent across the lifespan (Briley & Tucker-Drob, 2014), there is little work examining the extent to which polygenic effects associated with personality in adults are associated with personality in children and adolescents.

Exploratory approaches such as phenome-wide association studies (PheWAS; Bastarache et al., 2022), where associations of polygenic indices (PGIs) with measured phenotypes in a sample are estimated, can characterize how genome-wide genetic effects on adult personality are associated with behaviors, experiences, and outcomes, particularly in less studied contexts such as late childhood and early adolescence. Importantly, genotypes are set at conception, meaning PGIs act as stable indicators of the heritable aspects of a trait even in contexts (such as development) where phenotypic measurements may fluctuate (Allegrini et al., 2022). During periods of development where personality is rapidly shifting, such as childhood and adolescence, understanding behavioral correlates of genetic effects on personality may illuminate behaviors, environments, and life events that ultimately coalesce into stable personality traits (Belsky & Harden, 2019).

We conducted a PheWAS of polygenic indices for the Big 5 personality traits among 5,556 European-like ancestry (EUR) and 1,584 African-like ancestry (AFR) children at two timepoints (i.e., ages 9-11 and 11-13) in the Adolescent Brain Cognitive Development^SM^ Study (ABCD Study®) (Volkow et al., 2018). We performed cross-sectional and longitudinal analyses of phenotypes spanning cognition, screen time, demographics, substance use, environment, and physical and mental health domains to examine how polygenic variation associated with personality in adulthood is linked with thoughts, feelings, behaviors, and important outcomes in late childhood and early adolescence before the typical emergence of adult personality.

## Results

### Personality PGIs are associated with mental health, behavior, and environments in late childhood and early adolescence

We estimated associations between PGIs for adult Big 5 personality and a broad range of phenotypes (cognition, screen time, demographics, substance use, culture/environment, physical health, child mental health, family mental health) measured at ages 9-11 (n_phenotypes_=1,271) and 11-13 (n_phenotypes_=1,697) among EUR (n=5,556) and AFR (n=1,584) children. We used mixed effects regressions that included random intercepts for family and study site to account for nesting in ABCD, and age at assessment, sex, and the first 10 genetic principal components as covariates. We also used linear mixed effects regressions to conduct a PheWAS across imaging metrics in ABCD (Details in **Online Methods** and results reported in the **Supplementary Note**). We primarily report EUR results, as AFR results had limited power (**Online Methods**, detailed results reported in **Supplementary Note**).

Across the five domains, personality PGIs were associated with between 13 and 86 phenotypes at baseline (1%-5% of phenotypes; average magnitude of significant effects ranging from .07-.14; estimates for all PGIs with baseline phenotypes presented in **Supplementary Tables 1-5**) and 21 and 168 phenotypes at 2-year follow up (FU_2_; 2-10% of phenotypes; average magnitude of significant effects ranging from .06-.09; estimates for all PGIs with follow up phenotypes presented in **Supplementary Tables 6-10**); the neuroticism and openness PGIs were associated with the most and least phenotypes, respectively (**Figure 1**). Generally, personality PGIs were significantly associated with behaviors that are often indexed by adult personality measures (e.g., greater neuroticism PGI was associated with more worrying, greater extraversion PGI was associated with less shyness, and greater conscientiousness PGI was associated with getting things done on time). Personality PGIs were also associated with important health behaviors and outcomes, such as sleep problems (higher neuroticism PGI, lower conscientiousness PGI), use of mental health or substance use disorder services (lower agreeableness PGI, higher neuroticism PGI), greater sociability (higher agreeableness PGI, higher extraversion PGI) and better cognitive functioning (lower conscientiousness PGI, lower neuroticism PGI, higher openness PGI). Finally, personality PGIs were significantly associated with aspects of the home and family environment, such as greater family conflict (lower agreeableness PGI, higher neuroticism PGI) and more deprived neighborhood characteristics (higher neuroticism PGI).

**Figure 1.**
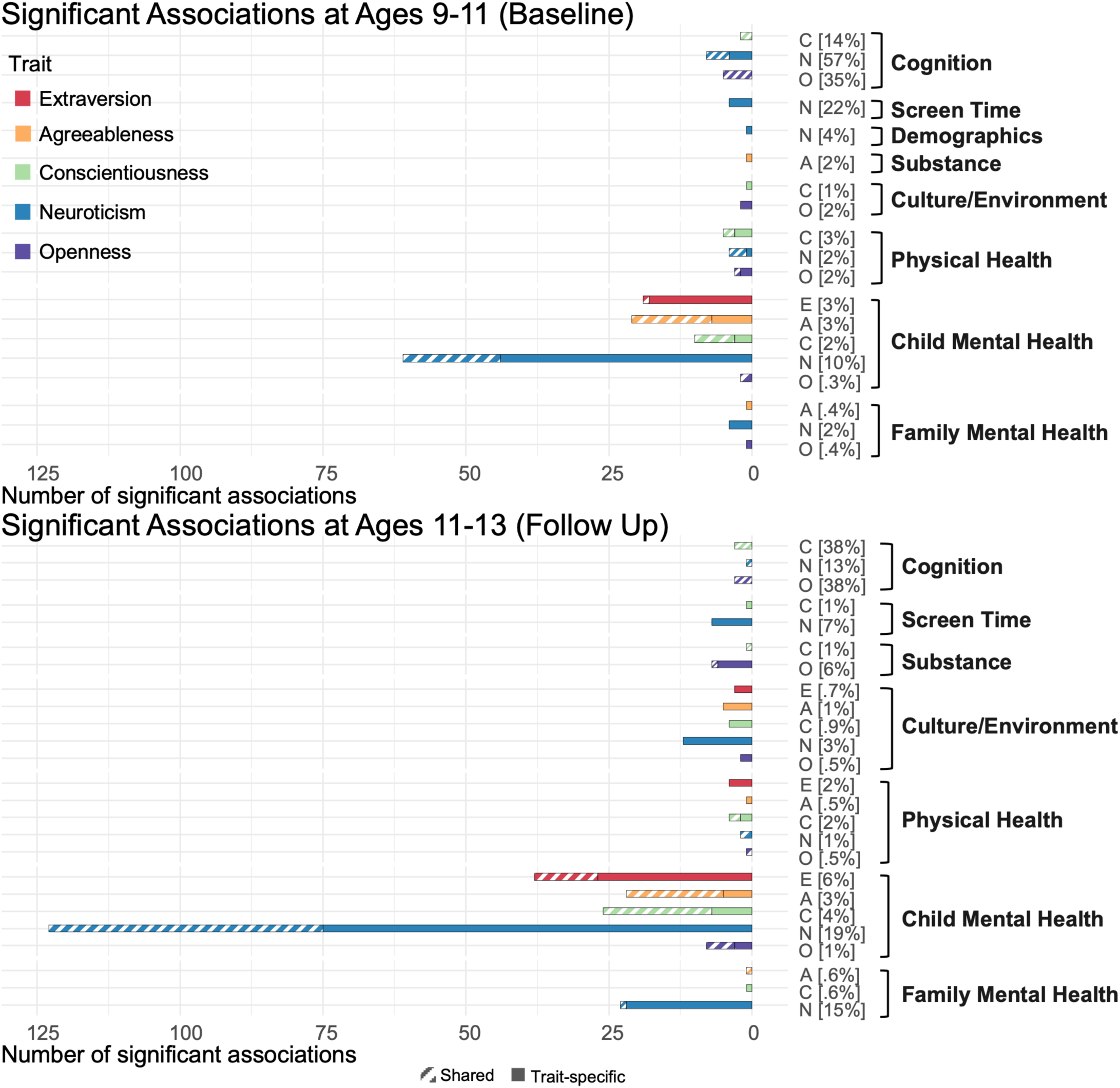
Bonferroni significant associations among baseline phenotypes (top), 2-year follow up phenotypes (bottom) and Big 5 PGIs. Solid colors are phenotype-PGI associations that are unique to a specific PGI, and striped bars represent phenotypes that are associated with more than one Big 5 PGI. Percentages represent percent of phenotypes in a domain significantly associated with the respective PGI. The majority of personality PGI associations are with child mental health phenotypes, particularly for the Neuroticism PGI. At 2-year follow up, phenotype associations are spread across more personality PGIs, e.g., the Conscientiousness PGI is associated with phenotypes in screen time, substance use, physical health, and child mental health.

There were no significant associations among personality PGIs and phenotypes in the AFR sample after multiple testing correction. However, the majority of effects in the AFR sample were similar in magnitude and direction to effects in the EUR sample; estimates in the AFR sample were accompanied by larger standard errors due to the smaller sample size (AFR sample estimates are presented in **Supplementary Tables 11-20**).

Greater genetic propensity to neuroticism was the most robust correlate of childhood mental health, including anxiety, depressed mood, and other internalizing symptoms, as well as endorsement of psychotic-like experiences (e.g., “I have seen things that other people apparently can’t see.”) at both timepoints (**Figure 2**). At baseline and FU_2_, higher neuroticism PGIs were associated with elevated likelihood of sleep disturbances, lower cognition scores, parental depression history, and lower parental educational attainment. The neuroticism PGI was also the only personality PGI related to increased screen time, with significant associations with multiple screen time items at both timepoints. At FU_2_, higher neuroticism PGI was also associated with poorer school performance and engagement, greater likelihood of parent divorce or separation, and experiencing more negative life events.

**Figure 2.**
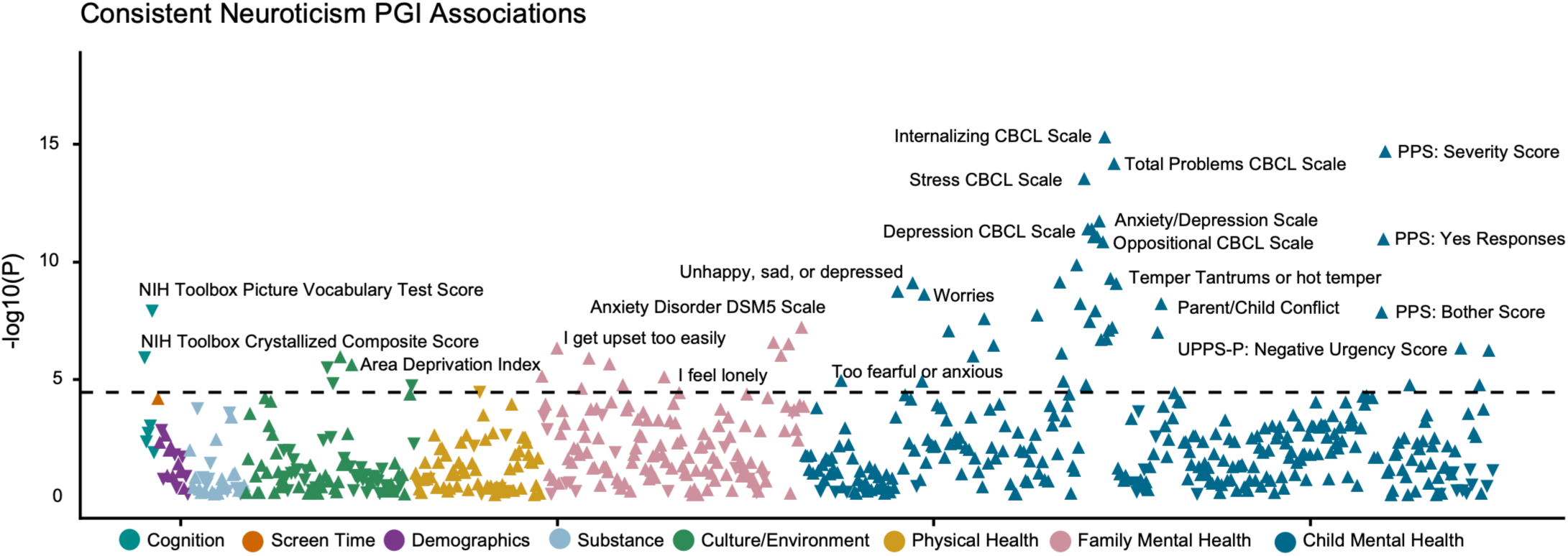
Bonferroni-significant associations at both baseline and FU2 between Neuroticism PGI and phenotypes. Triangles pointing up indicate positive PGI-phenotype associations, and triangles pointing down indicate negative PGI-phenotype associations. Dashed line signifies p-value threshold at which points are significant after Bonferroni correction for multiple testing. CBCL: Child Behavior Checklist, PPS: Prodromal Psychosis Scale, UPPS-P: UPPS-P Impulsive Behavior Scale.

The agreeableness PGI was also associated with childhood mental health, particularly externalizing-related behaviors at both timepoints (**Figure 3**). Youth with higher agreeableness PGIs were *less* likely to display aggressive, rule-breaking, and oppositional behaviors at baseline and FU_2_, and were less likely to receive outpatient services for mental health or substance use concerns at baseline. At FU_2_, higher agreeableness PGIs were associated with reduced family conflict, more prosocial behavior, and reporting living in a safer neighborhood. In supplementary analyses of imaging phenotypes, greater agreeableness PGIs were also associated with reduced head motion (Supplementary Table 29).

**Figure 3.**
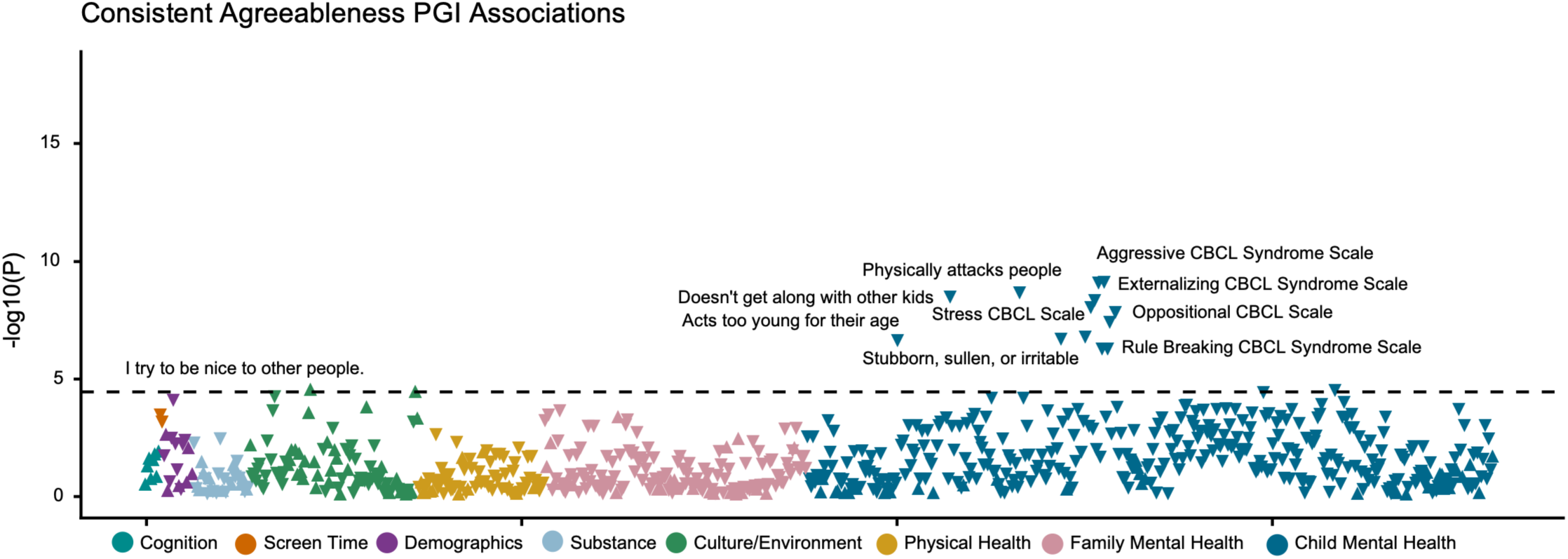
Bonferroni-significant associations at both baseline and FU2 between Agreeableness PGI and phenotypes. Triangles pointing up indicate positive PGI-phenotype associations, and triangles pointing down indicate negative PGI-phenotype associations. Dashed line signifies p-value threshold at which points are significant after Bonferroni correction for multiple testing. CBCL: Child Behavior Checklist.

At baseline and FU_2_, greater genetic propensity to extraversion was associated with variables reflecting sociability and assertiveness (e.g., “Talks too much” and “Bragging, boasting”), consistent with aspects of adult extraversion (**Figure 4**). Higher extraversion PGIs were also associated with reduced social anxiety symptoms at baseline. At FU_2_, higher extraversion PGIs were also significantly associated with greater surgency and physical activity and less shyness.

**Figure 4.**
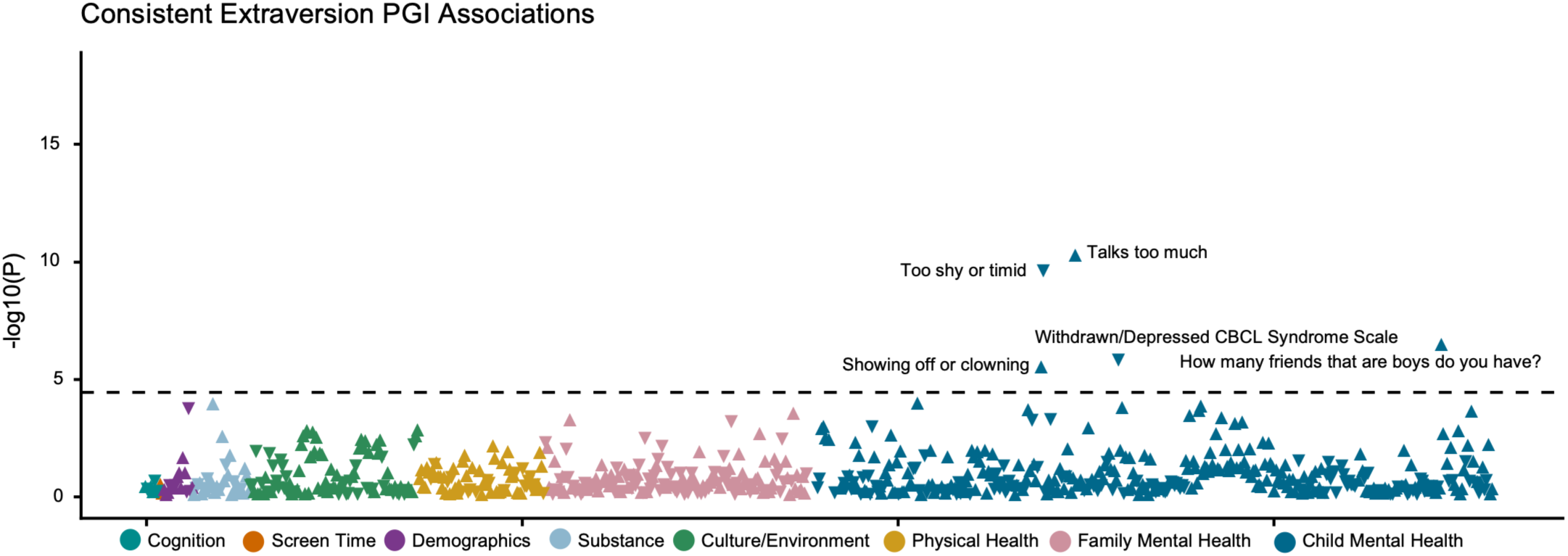
Bonferroni-significant associations at both baseline and FU2 between Extraversion PGI and phenotypes. Triangles pointing up indicate positive PGI-phenotype associations, and triangles pointing down indicate negative PGI-phenotype associations. Dashed line signifies p-value threshold at which points are significant after Bonferroni correction for multiple testing. CBCL: Child Behavior Checklist.

Genetic propensity to conscientiousness related predominantly to youth reports of being more organized (e.g., less likely to “often lose things”) and to fewer attention problems and greater perseverance (**Figure 5**). Higher conscientiousness PGIs also related to fewer sleep disturbances. In addition, the conscientiousness PGI was associated with measures of family organization (e.g. “Family members make sure their rooms are neat.”) at FU_2_. Interestingly, higher conscientiousness PGIs were associated with greater school engagement but also with lower crystallized cognition scores.

**Figure 5.**
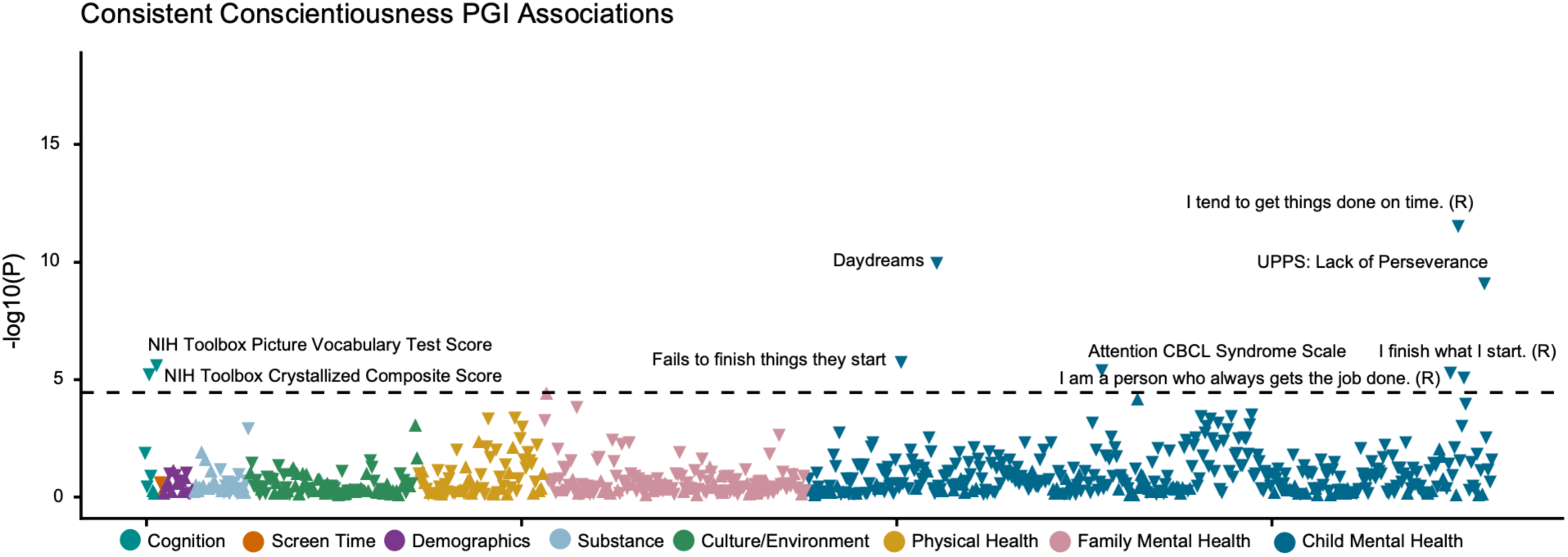
Bonferroni-significant associations at both baseline and FU2 between Conscientiousness PGI and phenotypes. Triangles pointing up indicate positive PGI-phenotype associations, and triangles pointing down indicate negative PGI-phenotype associations. Dashed line signifies p-value threshold at which points are significant after Bonferroni correction for multiple testing. (R) indicates questionnaires that are reverse coded. CBCL: Child Behavior Checklist, UPPS: UPPS-P Impulsive Behavior scale.

Genetic propensity to openness was associated with the fewest phenotypes at baseline and FU_2._ Higher openness PGIs were associated with better performance on crystallized cognitive measures (e.g., oral reading comprehension) and greater endorsement of daydreaming, consistent with openness including facets of both intellectual curiosity and fantasy (**Figure 6**). At FU_2_, the openness PGI was also associated with endorsing more negative expectancies for alcohol but more positive expectancies for cannabis. Finally, youth with higher genetic propensity to openness were more likely to identify as gay or bisexual (via both self- and parent-report).

**Figure 6.**
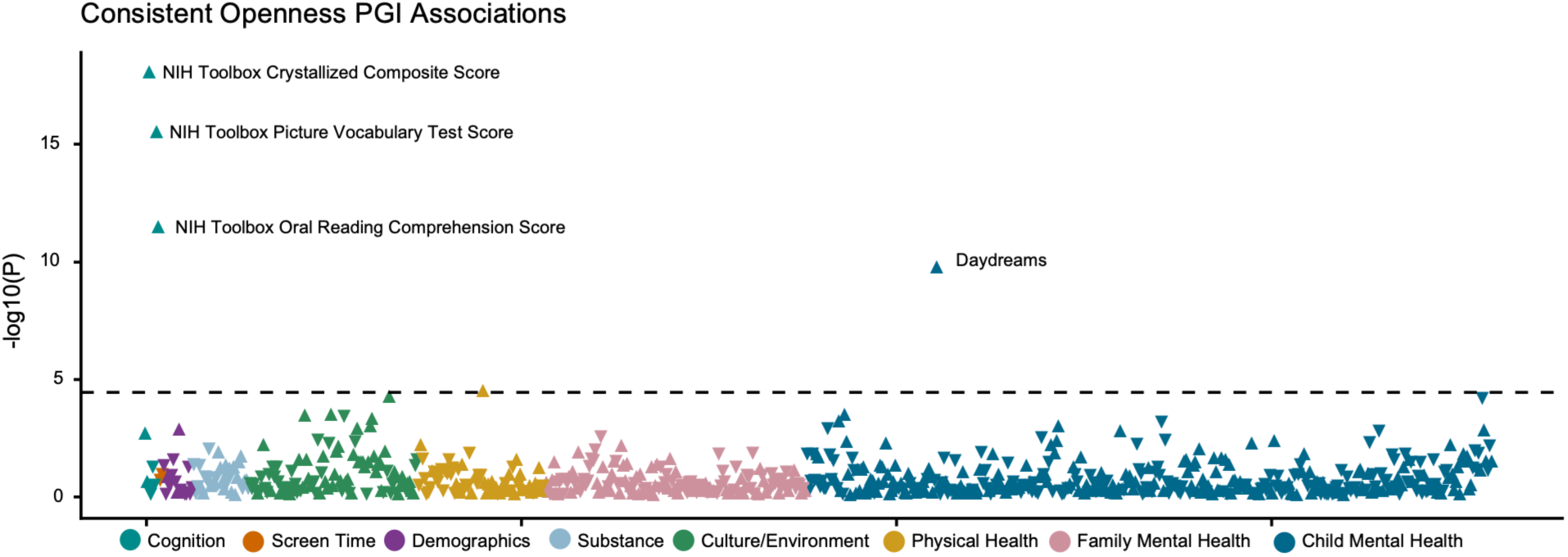
Bonferroni-significant associations at both baseline and FU2 between Openness PGI and phenotypes. Triangles pointing up indicate positive PGI-phenotype associations, and triangles pointing down indicate negative PGI-phenotype associations. Dashed line signifies p-value threshold at which points are significant after Bonferroni correction for multiple testing.

### Personality PGI associations shift between late childhood and early adolescence

We conducted multivariate analyses for 731 measures assessed at both timepoints to evaluate whether personality PGI associations changed between ages 9-11 and ages 11-13 (simultaneous regression estimates and likelihood ratio comparison tests for all PGIs and phenotypes presented in **Supplementary Tables 21-25**). Broadly, longitudinal analyses provided evidence for stability in personality PGI-phenotype associations by revealing largely consistent effects at baseline and FU_2_. However, likelihood ratio tests comparing effects between baseline and FU_2_ indicated several personality PGIs demonstrated stronger effects at FU_2_ relative to baseline or were associated with FU_2_ but not baseline variables; less frequently, effects were weaker at FU_2_ relative to baseline for the same variable.

The neuroticism, openness, and conscientiousness PGIs demonstrated stronger effects at FU_2_ relative to baseline for several variables reflecting aspects of each domain typically indexed by adult personality inventories. For example, the neuroticism PGI was more strongly associated with anxiety, depression, and stress symptoms; the openness PGI was more strongly associated with crystallized intelligence measures; and the conscientiousness PGI was more strongly associated with “finishing projects” at FU_2_.

Multiple personality PGIs also had stronger associations with school and social related traits at FU_2_. Higher neuroticism PGIs were associated with negative feelings towards school (e.g., not liking school, feeling less smart compared to other kids), and higher agreeableness PGIs were associated with more positive feelings towards school at FU_2_. Agreeableness and conscientiousness PGIs were more strongly associated with parent-reported prosocial behavior (e.g., “My child offers to help others” and “My child is helpful”) at FU_2_ than at baseline. This may reflect that, as children progress through school, positive social behavior is expressed more strongly at older ages.

In general, associations with cognitive test scores similarly strengthened between baseline and FU_2_. The conscientiousness PGI was more strongly associated with lower crystallized cognition scores and oral reading scores at FU_2_ than at baseline, and the openness PGI was more strongly associated with crystallized cognition scores at FU_2_. Conversely, the neuroticism PGI had a weaker association with Rey Auditory Verbal Learning Test scores at FU_2_.

Some behavioral associations with personality PGIs were only present at FU_2_, suggesting that some facets of personality domains may be expressed only at later ages. The extraversion PGI was associated with greater fun seeking and sensation seeking, and the agreeableness PGI was associated with being more likely to attend religious services only at FU_2_.

Personality PGI associations with substance use behaviors also shifted across baseline and FU_2_. Higher neuroticism and lower agreeableness PGIs were associated with greater caffeine consumption only at baseline, whereas higher conscientiousness PGIs were associated with being more curious about tobacco products only at FU_2_, likely reflecting that as children progress into late adolescence attitudes towards substance use are expressed as adolescents start to encounter and discuss substance use among peers.

### Within-family genetic variation in personality PGIs are associated with specific personality traits

We parsed significant personality PGI associations from our primary PheWAS into within- and between-family genetic effects in a subsample of EUR full-siblings (N = 1,842) (see **Methods;** Estimates presented in **Supplementary Tables 26-27**). Given the sibling structure of ABCD, within-family genetic effects were indexed using within-sibling PGIs, which reflect genetic differences between siblings that are free from shared/familial environmental influences and between-family confounding (e.g., population stratification). Significant within-family effects are inconsistent with passive gene-environment correlation effects where parents shape shared family environments based on their own genetic profiles, which are also shared with offspring.

The majority of PGI associations from our primary PheWAS did not have significant within-family effects, in part due to reduced power as a result of limiting the sample to siblings, reflected by increased standard errors for within and between-family PGI associations.

After accounting for between-family variation, within-family variation in the conscientiousness PGI was associated with being more likely to “finish things” (i.e., task completion) at FU_2_. Notably, within-family variation in the conscientiousness PGI was significantly associated with worse cognitive scores, while between-family variation was not significantly associated with cognition at baseline. Both within- and between-family variation in the openness PGI were significantly associated with more years of reading for pleasure, with larger effect sizes for the within-family PGI, suggesting that both the family environment and individual genetic predispositions impact reading behavior. Within-family genetic variation in the extraversion PGI was significantly associated with reporting feeling energized by being in large crowds of people, suggesting that child genetic factors may drive context specific manifestations of extraversion. Finally, within-family variation in the agreeableness PGI was associated with parents positively endorsing “There is very little I enjoy,” indicating that low child agreeableness PGIs were linked with negative parent emotions independently of parent genetic factors. There were no significant effects for the neuroticism PGI in within-family analyses.

## Discussion

Our findings document that adult personality genetics are associated with individual differences in youth behaviors and experiences, including early expressions of personality, potentially shaping the development of personality and other correlated characteristics. Overall, the findings are generally consistent with phenotypic associations observed between personality traits and outcomes in adulthood. Even though the ABCD Study did not include a youth assessment of the Big 5 personality traits, genetic liability to personality traits measured in adult cohorts demonstrated associations with personality markers such as worrying (neuroticism PGI), attention (conscientiousness PGI), and daydreaming (openness PGI) as early as age 9. Youth personality PGIs were also related to possible consequences of these attributes, such as larger peer groups (extraversion PGI), higher school engagement (conscientiousness PGI), and prosocial behavior (agreeableness PGI). These findings add to previous work suggesting that adult Big 5 personality traits can be identified in children and adolescents (Soto et al., 2008; Soto & Tackett, 2015; Tackett et al., 2012), specifically suggesting the phenotypic associations in childhood are early manifestations of adult personality genetics. Moreover, these childhood associations establish the convergent and predictive validity of the adult personality PGI, substantially bolstering confidence in its capacity to capture genetic propensities across the lifespan.

Mirroring well-established associations between normal range personality measures and clinical outcomes, polygenic propensity to adult personality was associated with a broad and clinically meaningful range of youth behaviors. The neuroticism PGI was most closely related to child mental health, especially internalizing symptoms and disrupted sleep. Notably, higher neuroticism PGIs were linked to increased screen time. While emerging studies of this controversial and actionable youth behavior have linked it to subsequent anxiety and depressed mood (Nagata et al., 2024, 2025), our findings suggest that pre-existing vulnerability to neuroticism (and related traits such as internalizing behavior) may contribute to excess engagement with screens, which may then further isolate youth and exacerbate risk (Zhang et al., 2023).

Youth externalizing behaviors were primarily related to lower genetic propensity for agreeableness. Lower agreeableness PGIs were associated with higher scores of conduct problems, aggression, and attention problems, consistent with phenotypic associations in adults (Miller et al., 2008). Higher extraversion PGIs and lower conscientiousness PGIs were also related to aspects of youth externalizing (impulsivity and inattention, respectively), though neither PGI was significantly associated with clinical externalizing scales or conduct problems. These results are consistent with links observed in adults between extraversion and lower conscientiousness and more impulsive or inattentive aspects of externalizing in particular (Creswell et al., 2019).

Cognition and academic achievement were most closely related to conscientiousness and openness PGIs (Malanchini et al., 2019; Tucker-Drob et al., 2016), albeit with a seemingly paradoxical association between higher conscientiousness PGIs and lower crystallized intelligence scores. The negative association between the conscientiousness PGI and these cognitive scores likely reflects sampling bias in the original conscientiousness GWAS, which is negatively genetically correlated with educational attainment (Schwaba et al., 2026). Generally, large biobank and other large-scale samples used in GWAS report greater educational attainment than the general population (Fry et al., 2017; Schoeler et al., 2023; Tyrrell et al., 2021), and in highly educated samples, conscientiousness is often negatively correlated with cognitive function due to college selection (Murray et al., 2014). The paradoxical association of conscientiousness PGIs with greater school engagement and worse cognitive scores in late childhood may alternatively reflect that youth with greater conscientiousness are more likely to overcome initial cognitive deficits to achieve academic success, consistent with the compensatory model of cognition and conscientiousness interplay (Hufer-Thamm et al., 2023; O’Connell & Marks, 2022). Future work studying the relationship among genetic propensity for conscientiousness, childhood traits, and academic outcomes across development may offer more insight.

Correlates of the Big 5 PGIs were broadly consistent across the two-year follow-up period (i.e., baseline to FU_2_); however, there was also support for emerging phenotypic correlates of extraversion and agreeableness, such as sensation seeking and prosocial behavior (respectively). As individuals mature, facets such as conscientiousness and agreeableness might emerge and develop concurrently (DeYoung, 2006; Soto & Tackett, 2015), while decreases in traits and behaviors typical of extraversion are also common in adulthood and later life (Bleidorn et al., 2022; Roberts et al., 2006). The transition from baseline to FU_2_ also represents entry into middle school, a period marked by pubertal shifts and changing social structures, enhancing extraversion’s salience (Brook & Schmidt, 2020; Roberts et al., 2006). Future analyses of the Big 5 PGIs with longitudinal trends in behavior and experience could provide insights into behavioral maturation in adolescent cohorts.

Some limitations are noteworthy. GWAS and, by extension, PGIs can be confounded by population stratification, assortative mating, and patterns of gene-environment correlation (Brumpton et al., 2020; Veller & Coop, 2024). We incorporated within-sibling PGI analyses to address these potential sources of bias. We did not find many significant within-sibling associations. However, these analyses were less well-powered than analyses in the full sample, and previous within-family analyses of the Big 5 personality traits indicate personality may be less confounded than other phenotypes (Schwaba et al., 2026). Additionally, we performed analyses in AFR individuals, however, these analyses were underpowered due to small AFR GWAS samples. Despite limitations in statistical power, the majority of effect estimates in the AFR sample were of similar magnitude and direction as those estimated in the EUR sample.

We show that personality genetics are associated with a variety of youth behaviors linked with achievement, psychosocial adjustment, mental health, and treatment engagement. Understanding the short- and long-term sequelae of genetic propensity to specific personality traits might provide important insights into mechanisms of personality development throughout adolescence and identify individuals in need of additional support for mental health, academic achievement, and social functioning.

## Online Methods

### Sample and Measures

#### Sample

The Adolescent Brain Cognitive Development (ABCD) Study is an ongoing longitudinal study of child/adolescent development across 21 research sites in the United States (Volkow et al., 2018). Our analyses were restricted to individuals with European-like genomic ancestry (EUR) that completed the baseline (n=5,556; 9-11 years of age; 47% female) and 2-year follow-up assessment (n=5,048; 11-13 years of age; 47% female). We primarily report results from EUR-like individuals (AFR-like ancestry individual results reported in the **Supplementary Note**), due to the lack of well-powered GWASs in other ancestral populations, challenges porting GWAS summary statistics across broad ancestry groups to estimate polygenic indices, as well as small groups of other ancestries in ABCD (Duncan et al., 2019; Martin et al., 2019; Wang et al., 2020). Parental/caregiver written and child verbal informed consent/assent were provided for a research protocol approved by a centralized institutional review board as well as institutional review boards at each data collection site (https://abcdstudy.org/sites/abcd-sites.html).

#### Measures

Phenotypes were selected from a broad sample of variables related to substance use, mental health, physical health, cognition, culture and environment, demographics, and screen time that were measured at baseline and 2-year follow up visit. Genetic data were drawn from release 3.0, and phenotypic data was drawn from release 4.0. PheWAS item selection is described in detail in Paul et al. (2024). Administrative, redundant, and irrelevant items (e.g., number of missing items) were removed. Variables with fewer than 100 data points or fewer than 100 endorsements of minority categories of categorical variables were also excluded. 621 phenotypes at baseline and 454 phenotypes at 2-year follow up were categorical items that were dummy coded, (i.e., diagnosis codes or symptom endorsements). Additional details on phenotype coding and selection processes are described in Paul et al. (2024). We used a final selection of 1,271 phenotypes at baseline and 1,697 at follow up.

#### Polygenic Indices

Genome-wide association summary (GWAS) statistics for Extraversion, Agreeableness, Conscientiousness, Neuroticism, and Openness were used from the largest available GWAS of the Big 5 domains (Schwaba et al., 2026). We derived weights for polygenic indices (PGIs) using SBayesRC (Zheng et al., 2024), a Bayesian method that incorporates functional annotation data to distinguish likely causal SNPs from non-causal SNPs. SBayesRC uses a low-rank model to model the LD among common variants, then uses a multicomponent annotation-dependent mixture prior to model the distribution of SNP effect sizes. For all PGIs, we used probability starting values of 95%, 2%, 2%, and 1%, with effect sizes scaled at .00, .01, .10, and 1.00. We used BaselineLD v2.2 genomic annotation data (Gazal et al., 2017) and approximately 7 million imputed common SNPs from UK Biobank for LD reference (Zheng et al., 2024). We excluded the MHC region from analysis. PLINK2 (Chang et al., 2015) was used to estimate PGIs for each European-like ancestry ABCD participant using the SBayesRC weights. ABCD genetic data imputation and quality control measures are described in detail in Fan et al. (2023) and Paul et al. (2024). Briefly, the Rapid Imputation and COmputation PIpeLIne for GWAS (RICOPILI) was used to perform QC on individuals with ABCD phase 3.0 genetic data using default parameters (genotyping call rate > 98%, inbreeding coefficient (F) < +-0.2, and sex checks) (Lam et al., 2020). Variants with missingness > 2% and Hardy-Weinberg equilibrium *p* < 1x10^-6^ were excluded. Genetic ancestry was confirmed by performing principal component analysis in RICOPILI with EIGENSTRAT15 using 1000 Genomes European and African reference populations. Only SNPs with Rsq > 0.8 and minor allele frequency > 0.01 were retained for polygenic scoring.

### Statistical Analyses

#### Cross-sectional Phenome-wide Association Analyses of Big 5 PGIs at Baseline and Follow-up

We used mixed effects regression models to test associations between PGIs for each Big 5 domain and phenotypes at baseline and 2-year follow up. Continuous variables were standardized prior to analysis. We used linear mixed effects models for continuous variables and generalized linear mixed effects models for dichotomous outcomes using the lme4 package in R. All models included age at assessment, sex, and the first 10 genetic principal components as covariates and included random intercepts for family and study site.

Analyses of phenotypes assessed at 2-year follow up also included a covariate indicating whether the visit was completed in person, remotely, or a hybrid due to the COVID-19 pandemic. To adjust for the number of tests in non-imaging phenotypes (1,271 baseline, 1,690 follow up), we used Bonferroni correction for the set of models within each PGI (e.g., we corrected p-values for 1,271 tests for the neuroticism PGI in the baseline data). For analyses of brain imaging data (254 structural imaging metrics, 76 diffusion MRI metrics, 91 resting state MRI metrics; results provided in **Supplementary Note**) we corrected within data subgroups (e.g., for the results of cortical imaging phenotypes, for each PGI, we corrected for the number of cortical imaging tests, rather than correcting for all tests across all imaging data). Additional details about analyses of the imaging data are presented in the supplementary note.

#### Comparing strength of PGI associations at baseline and 2-year follow up

We evaluated whether phenome-wide PGI associations varied across baseline and follow up for 731 measures assessed at both timepoints. In a multivariate model, phenotype measures at baseline and FU_2_ were simultaneously regressed on the personality PGI, and the covariance between baseline and FU_2_ measures was estimated. This approach accounts for the stability in repeated measurements over time, indexing the effect of the PGI on the measurement outcome at time 2 while allowing residuals to correlate between measurement at time 1 and measurement at time 2.

Timepoint comparison analyses were performed using MPlus version 8.0 (Muthén & Muthén, 1998-2017). We ran two sets of models for each phenotype; one model in which the effect of the PGI was freely estimated across both timepoints, and one in which the effect of the PGI on the phenotype was constrained to be equal across both timepoints. We conducted model difference tests to assess whether the models where the PGI effect on outcomes was the same at both timepoints significantly differed from models where the PGI effect varied across time. Models for continuous outcomes used robust maximum likelihood estimation and we conducted model difference tests using Satorra-Bentler likelihood ratio tests (Satorra & Bentler, 2010). Models with categorical outcomes used logistic regression models with mean and variance adjusted weighted least squares estimation, and model difference testing was conducted using scaled chi-square difference tests through the Mplus DIFFTEST function. All models accounted for nesting of participants within data collection sites (stratified by site factor) and family relatedness (clustered by family ID variable). We included sex, baseline age, and the first 10 genetic principal components as covariates in all models. We used Bonferroni correction to adjust for the 731 tests conducted for each PGI.

#### Within-Sibling Analyses

We performed post-hoc within-sibling analyses to examine whether personality PGI effects on outcomes plausibly represent direct genetic effects. Direct genetic effects are the effects of an individual’s genetic variants on that same individual’s traits or behavior. Polygenic score associations can be confounded by three types of gene-environment correlation (rGE): passive rGE, where parents shape a child’s environment based on their own genetic predispositions; active rGE, where children select environments the correlate with their genetic predispositions; and evocative rGE, where a child evokes responses that correlate with their genetic predispositions (Jaffee & Price, 2007). Within-sibling analyses test whether PGI associations are inconsistent with passive rGE, though evocative and active rGE still influence within-sibling variation in genetic effects (Brumpton et al., 2020; Howe et al., 2022; Kong et al., 2018).

Within-sibling analyses were conducted in a subsample of siblings (*n* = 1842) from our original EUR-like ancestry sample. We computed mean family PGIs, which are average polygenic indices across all family members, and individual PGIs, which are each sibling’s deviation from their family mean PGI. Mean family and individual PGIs were included as predictors in a mixed effects model, with age at assessment, sex, and the first 10 PCs included as covariates. Models also included random intercepts for family and study sites. We estimated within-sibling models only for outcomes that were significantly associated with personality PGIs in our primary analysis at baseline or follow up, for a total of 155 measures across all 5 PGIs at baseline (19 for extraversion PGI, 23 for agreeableness PGI, 18 for conscientiousness PGI, 82 for neuroticism PGI, 13 for openness PGI) and 302 outcomes across all 5 PGIs at follow up (45 for extraversion PGI, 28 for agreeableness PGI, 40 for conscientiousness PGI, 168 for neuroticism PGI, 21 for openness PGI).

## Data Availability

All data produced in the present study are available upon reasonable request to the authors.

## Acknowledgements

The authors acknowledge the Research Infrastructure Services (RIS) group at Washington University in St. Louis for providing computational resources and services needed to generate the research results delivered within this paper. This work also used the High Throughput Computing Facility at the Center for Genome Sciences and Systems Biology at Washington University in St. Louis.

