## Supplementary Note for "Childhood Correlates of Genetic Propensity to Personality Traits"

*PheWAS of Imaging Phenotypes in ABCD*

A detailed description of imaging acquisition, processing, and quality control procedures are detailed in Casey et al., 2018. T1-weighted structural images were obtained via 3T MRI scanners. Real-time motion detection and correction was implemented to mitigate the influence of head motion. Full QC procedures for processed imaging data are described in Hagler et al (2019). Imaging derived phenotypes from structural MRI (sMRI), resting state fMRI (rs-MRI), and diffusion MRI (dMRI) were included for a total of 421 neuroimaging phenotypes.

MRI data were pre-processed using FreeSurfer and images were registered to the Desikan atlas (Hagler et al., 2019). Only MRI data that passed FreeSurfer QC criteria were retained (Additional details in Paul et al., 2024). sMRI imaging derived phenotypes included 15 global brain metrics and 239 regional sMRI metrics spanning cortical and subcortical gray matter volumes, thicknesses, and surface areas. dMRI phenotypes were derived from mean diffusivity (MD) and fractional anisotropy (FA) metrics, which were calculated with a linear estimation approach with log-transformed, diffusion-weighted signals. AtlasTrack was used to label major white matter tracks (Hagler Jr. et al., 2009). dMRI phenotypes included a global FA measure, 37 regional FA measures, a global MD measure, and 37 regional MD measures. rs-MRI phenotypes were derived from pairwise correlations among regions of interest within functionally defined parcellations (Gordon networks: Gordon et al., 2016). rs-MRI phenotypes were Fisher Z-transformed correlation values within the 13 Gordon networks across 78 additional between-network rs-fMRI metrics (91 phenotypes total).

Imaging data was available for 5,556 EUR children. Linear mixed effects models were used to test associations between PGIs of the Big 5 domains and imaging phenotypes. Imaging variables were standardized prior to analysis. Models of imaging phenotypes included age at assessment, sex, scanner type and average head motion (for rs-MRI and dMRI models) as covariates. All models included random intercepts for family and study site. Global surface area was included as a covariate for regional surface area models, mean thickness was included as a covariate in regional thickness measures, and global volume was included as a covariate in regional volume measures. Total fractional anisotropy (FA) across all fibers was included as a covariate in models with FA measuers and mean diffusivity was included as a covariate in diffusivity models. Bonferroni correction was applied within imaging modalities (e.g. dMRI, cortical area measures, global measures, etc.) and within PGIs.

Regression estimates of the effect of each big 5 PGI on imaging phenotypes are presented in Supplementary Tables 28-32. The agreeableness PGI was significantly associated with reduced head motion in models of dMRI phenotypes. The openness PGI was significantly associated with greater global subcortical volume in sMRI analyses. In dMRI analyses, the openness PGI was significantly associated with greater fractional anisotropy in the left inferior longitudinal fasciculus, and the agreeableness PGI was significantly associated with reduced average fractional anisotropy in the forceps major. Of the rs-MRI phenotypes, the agreeableness PGI was significantly associated with reduced correlation between cingulo-opercular and sensorimotor mouth networks, and the neuroticism PGI was associated with more strongly correlated auditory and visual networks.

*PheWAS of participants with African-like ancestry*

We conducted an additional PheWAS of the big 5 PGIs in ABCD participants with African-like ancestry. Available GWAS summary data for the big 5 personality traits was limited for AFR ancestry groups (AFR N ~ 43,000 compared to EUR N ~ 650,000), meaning PGIs generated from available AFR data have reduced power to predict personality than those generated from EUR data (Wang et al., 2022). Schwaba et al. (2026) showed that personality PGIs were more predictive in AFR-like populations using weights generated from EUR summary statistics, rather than using methods that combine EUR and AFR data (e.g., PRS-CSx). We used SBayesRC (Zheng et al., 2024), a Bayesian method that incorporates functional annotation data to distinguish likely causal SNPs from non-causal SNPs, to derive PGI weights from EUR GWAS summary data (additional details in **Online Methods**) and estimated PGIs for each AFR-like ABCD participant (N = 1,584) using PLINK2 (Chang et al., 2015). Although PGIs generated from EUR summary data are better able to predict personality due to greater sample size, PGIs have limited cross-ancestry portability, resulting in reduced power to detect significant associations between phenotypes and PGIs in AFR target samples (Wang et al., 2022).

Mixed effects regression models were used to test associations between PGIs and phenotypes at baseline ($n_{phenotypes}$ = 940) and 2-year follow up ($n_{phenotypes}$ = 1,351) in 1,584 AFR ancestry children. Fewer phenotypes were available at baseline and follow-up for the AFR group due to reduced sample size, increasing the number of variables that had to be removed due to insufficient item endorsement (for additional information see Paul et al., 2024). We used the same mixed effects regression models, covariates, and significance correction methods that were used in the EUR sample (**Online Methods**). Patterns of associations among the big 5 PGIs and phenotypes were similar to those in the EUR sample across both timepoints, but very few associations were statistically significant in the AFR sample due to reduced power. At baseline, the openness PGI was significantly associated with greater scores on the WISC matrix reasoning test. No other PGI-baseline phenotype associations were significant after Bonferroni correction. 83-90% of EUR PGI-phenotype effect estimates were within the 95% confidence interval of the AFR PGI-phenotype estimates, indicating that effect sizes were largely similar between groups and that differences in statistical significance were largely a result of reduced power in the AFR group. At follow-up, the openness PGI was significantly associated with greater scores on the NIH toolbox picture vocabulary test. The agreeableness PGI was significantly associated with often interrupting others after FDR correction at follow up, however, this association was nonsignificant with Bonferroni correction. 84-90% of EUR PGI-phenotype effect estimates were in the 95% confidence interval of the AFR PGI-phenotype estimates at follow-up.
